# Early Spread of SARS-Cov-2 in the Icelandic Population

**DOI:** 10.1101/2020.03.26.20044446

**Authors:** Daniel F Gudbjartsson, Agnar Helgason, Hakon Jonsson, Olafur T Magnusson, Pall Melsted, Gudmundur L Norddahl, Jona Saemundsdottir, Asgeir Sigurdsson, Patrick Sulem, Arna B Agustsdottir, Berglind Eiriksdottir, Run Fridriksdottir, Elisabet E Gardarsdottir, Gudmundur Georgsson, Olafia S Gretarsdottir, Kjartan R Gudmundsson, Thora R Gunnarsdottir, Arnaldur Gylfason, Hilma Holm, Brynjar O Jensson, Aslaug Jonasdottir, Frosti Jonsson, Kamilla S Josefsdottir, Thordur Kristjansson, Droplaug N Magnusdottir, Louise le Roux, Gudrun Sigmundsdottir, Gardar Sveinbjornsson, Kristin E Sveinsdottir, Maney Sveinsdottir, Emil A Thorarensen, Bjarni Thorbjornsson, Arthur Löve, Gisli Masson, Ingileif Jonsdottir, Alma Moller, Thorolfur Gudnason, Karl G Kristinsson, Unnur Thorsteinsdottir, Kari Stefansson

**Affiliations:** deCODE genetics/Amgen Inc., Reykjavik, Iceland; School of Engineering and Natural Sciences, University of Iceland, Reykjavik, Iceland; Department of Anthropology, University of Iceland, Reykjavik, Iceland; Department of Clinical Microbiology, Landspitali University Hospital, Reykjavík, Iceland; Directorate of health, Reykjavik, Iceland; Faculty of Medicine, School of Health Sciences, University of Iceland, Reykjavik, Iceland; BioMedical Centre of the University of Iceland, Reykjavik, Iceland

## Abstract

**BACKGROUND:** Limited data exist on how SARS-CoV-2 enters and spreads in the general population.

**METHODS:** We used two strategies for SARS-CoV-2 testing: targeted testing of high-risk individuals (n=4,551) and a population screening (n=5,502). We sequenced SARS-CoV-2 from 340 individuals.

**RESULTS:** On March 22 2020, 528 had tested positive for SARS-CoV-2 in the targeted testing (11.6%) and 50 in the population screening (0.9%); approximately 0.2% of the Icelandic population. Large fractions of positives had travelled outside Iceland (38.4% and 34.0%). Fewer under 10 years old were positive than those older: 2.8% vs. 12.3% for targeted testing (P=1.6e-9) and 0.0% vs. 1.0% for population screening (P=0.031). Fewer females were positive in the targeted testing than males (9.5% vs. 14.6%, P=6.8e-9). SARS-CoV-2 came from eight clades, seven A clades and one B clade. The clade composition differed between the testing groups and changed with time. In the early targeted testing, 65.0% of clades were A2a1 and A2a2 derived from Italian and Austrian skiing areas, but in the later targeted testing went down to 30.6% and were overtaken by A1a and A2a, the most common clades in the population screening.

**CONCLUSION:** SARS-CoV-2 has spread widely in Iceland outside of the high-risk groups. Several strains cause these infections and their relative contribution changed rapidly. Children and females are less vulnerable than adults and males. To contain the pandemic we must increase the scope of the testing.

## Introduction

Severe acute respiratory syndrome coronavirus 2 (SARS-CoV-2) caused clusters of severe respiratory illness in Wuhan, China in late 2019^1^. It was isolated and sequenced by January 2020 ^2,3^. As of March 23 2020, 332,935 individuals in 190 countries had been reported to be positive for SARS-CoV-2. SARS-CoV-2 causes COVID-19, a disease that has caused of 14,510 deaths^4^. As the number of new cases has decreased drastically in China, it has rapidly increased in Europe and the USA with the total number of COVID-19 associated deaths in Italy exceeding that in China^5^. By March 11 2020, The World Health Organization (WHO) made the assessment that COVID-19 should be characterized as a pandemic^6^. At great economic cost, many countries have adopted unprecedented measures to curb the spread of the virus such as large-scale use of isolation and quarantine, closing their borders, limits of public gathering and nation-wide lockdowns.

Early reports on COVID-19 case series from China and Italy indicate that SARS-CoV-2 causes a varying degree of illness^7,8^ with females and children being underrepresented among cases, in particular among severe and fatal cases. It is unclear if this is because females and children are less likely to be infected by SARS-CoV-2 or to develop COVID-19.

The first SARS-CoV-2 infection in Iceland was confirmed on February 28 2020 and by March 23, 588 individuals had been tested positive^9^. Iceland is an island with 364,000 inhabitants, with only one major gateway into the country, the international airport with 7 million passengers per year.

Here, we show how SARS-CoV-2 entered Iceland and began to spread within the country. We used two strategies for SARS-CoV-2 testing in Iceland, targeted testing of individuals at high risk of infection and a population screening of those who accepted an offer of free testing.

The genome of SARS-CoV-2 has been sequenced^2^, revealing close relationship to corona viruses such as SARS-CoV-1^10,11^ and MERS^12^ and researchers have deposited 1034 SARS-CoV-2 sequences in GISAID^13^. We sequenced the genomes of SARS-CoV-2 from those who tested positive to establish the origin of the virus and how it spreads in the population.

## Methods

### SARS-COV-2 TESTING

SARS-CoV-2 infected individuals were identified in Iceland through both targeted testing of high-risk individuals and population screening. The targeted testing started on January 31 2020, and has focused on individuals at high risk of infection because they were symptomatic (cough, fever, aches, and shortness of breath) or coming from ski areas in Austria and northern Italy, their contacts and those who contacted the healthcare system. As of March 19, the high-risk areas designation was extended to the whole world outside Iceland. As of March 22, 4,551 individuals had been tested in this targeted screening. The population screening for SARS-CoV-2 was initiated by deCODE on March 14 when 323 individuals had tested positive for SARS-CoV-2 by the targeted screening. The screening was open to everyone symptom free or with mild symptoms of common cold that is highly prevalent in Iceland at this time of the year. The registration for the test was online and during sample collection recent travels, contacts with positive individuals, and symptoms compatible with COVID-19 were registered. Over a six-day period 5,502 Icelanders were tested for SARS-CoV-2. All newly positives were placed in isolation and those who had been in contact with the positives, in quarantine for 2 weeks. All symptomatic individuals in quarantine were also tested. In addition to isolating positives and quarantining those at high risk of infection, on March 16 the Icelandic authorities initiated a ban on mass gathering above 100 people and stated that a social distancing of at least 2 meters should be maintained. On March 24, the ban on mass gathering was extended to 20 people. For SARS-CoV-2 testing, it was recommended to take both nasopharyngeal and oropharyngeal samples. RNA from all samples was isolated within 24 hours.

### TRACKING OF SARS-COV-2 INFECTIONS

All individuals who tested positive for SARS-CoV-2 were contacted by phone by a team designated by the authorities to track their infection. They were asked about their symptoms and when they started, recent travels and previous contacts with infected individuals. They were also asked to identify everyone whom they had been in contact with 24 hours before noticing their first symptom and for how long they interacted with each individual and how intimate the interaction was. All registered contacts were contacted by phone, requested to go into 2 weeks quarantine and asked about symptoms. Those with symptoms and those who developed them in quarantine were tested for SARS-CoV-2.

### RNA EXTRACTION

Viral RNA samples were extracted either at the Department of Clinical Microbiology laboratory at Landspitali, the National University Hospital of Iceland (LUH) or at deCODE. Both extraction methods are based on an automated magnetic bead-purification procedure, which includes cell lysis and Proteinase K treatment. RNA from samples at LUH were extracted (32 samples per 60 min run) using the MagNA Pure LC 2.0 instrument from Roche LifeScience, with 200/100 µL input/output volume(s), respectively. Samples at deCODE were extracted from swabs (96 samples per 70 min run) using the Chemagic Viral RNA kit on the Chemagic360 instrument from Perkin Elmer, with 300/100 µL input/output volume(s), respectively. Each step in the workflow was monitored using an in-house LIMS (VirLab) with 2D barcoding (Greiner, 300 µL tubes) of all extracted samples.

### TESTING OF SAMPLES FOR SARS-COV-2

Testing for SARS-CoV-2 was performed either at LUH or deCODE using similar quantitative real-time PCR (qRT-PCR) methods. TaqMan™ Fast Virus 1-step Master Mix, 2019-nCoV Assay kits v1 and 2019-nCov control kits were obtained from Thermo Fisher. Assay mix A, B and C were prepared containing FAM™ dye labelled probes for the SARS-CoV-2 specific genes ORF1ab, S-protein and N-protein, respectively. In addition, each assay mix contained VIC™ dye labelled probes for human RNase P as internal control. Samples from 96-well RNA sample plate(s) were dispensed into three wells each in a 384 plate layout, in addition to three negative (no template) and three positive controls. Assay mix was added in a total reaction volume of 12.5 µL per sample. All sample aliquoting and mixing at deCODE was performed with an automated Hamilton STARlet 8-channel liquid handler and the assay plates were scanned in an ABI 7900 HT RT-PCR system following manufacturer’s instructions with a total of 40 cycles of amplification. Samples with FAM™ dye C_t_ values <37 in at least two of three assays were classified as positive. Samples with FAM™ dye C_t_ values between 37 and 40 were classified as inconclusive and their testing repeated. If repeated testing gave the same result the sample was classified as positive. Samples with undetected FAM™ dye C_t_ values or values equal to 40 in all three assays were classified as negative if the human RNaseP assay was positive (VIC™ dye C_t_ <40). Validation of the RNA extraction and the qRT-PCR method(s) at deCODE was performed using 124 samples that had previously tested positive (n=104) or negative (n=20) with the qRT-PCR assay at LUH. All of the negative samples tested negative at deCODE and 102 of the 104 positive tested at LUH were also positive at deCODE. Two samples that tested positive at LUH were negative at deCODE. Upon subsequent sequencing (see below) viral genome could not be detected in these samples, probably because very few viral particles were present.

### STATISTICAL ANALYSIS

We used Fisher’s exact test to test for differences in frequency of positives below (or equal to) 10 years of age compared to older than 10 years of age and above 80 years of age compared to below (or equal to) 80 years of age. We used a likelihood ratio test based on logistic regression to calculate P values for differences in frequency of positives between sexes, using age and age squared as covariates. We used variance estimates based on binomial distribution to calculate 95% confidence intervals of fractions of positives.

### SAMPLE PREPARATION FOR SEQUENCING

Reverse transcription (RT) and multiplex PCR was performed based on information provided by the Artic Network initiative (https://artic.network/) to generate cDNA. In short, extracted viral RNA was pre-incubated at 65 °C for 5 min in the presence of random hexamers (2.5 µM) and dNTP’s (500 µM). Sample cooling on ice was then followed by RT using SuperScript IV (ThermoFisher) in the presence of DTT (5 mM) and RNaseOUT inhibitor (Thermo Fisher) for 10 min at 42°C, followed by 10 min at 70 °C. Multiplex PCR of the resulting SARS-CoV-2 cDNA was performed using a tiling scheme of primers, designed to generate overlapping amplicons of approximately 800 bp (Table S1). The primers were generously provided by Dr. David Stoddard at Oxford Nanopore Technologies. Two PCR reactions were done for each sample using primer pools A and B, respectively (Table S1). PCR amplification was done using the Q5^®^ Hot Start High-Fidelity polymerase (New England Biolabs) with primers at 1 µM concentration. The reactions were performed in an MJR thermal cycler with a heated lid at 105 °C, using 35 cycles of denaturation (15 sec at 98 °C) and annealing/extension (5 min at 65 °C). The resulting PCR amplicons were purified using Ampure XP magnetic beads (Beckman Coulter) and quantified using the Quant-iT™ PicoGreen dsDNA assay kit (Thermo Fisher). Amplified samples (20-500 ng) were randomly sheared using focused acoustics in 96-well AFA-TUBE-TPX plates (Covaris Inc.) on the Covaris LE220plus instrument with the following settings: Sample volume, 50 µL; temperature, 10 °C; peak incident power, 200W; duty factor, 25%; cycles per burst, 50; time, 350 sec. Sequencing libraries were prepared in the 96 well Covaris plates, using the NEBNext^®^ Ultra II kit (New England Biolabs) following the manufacturer’s instructions. In short, end repair and A-tailing was performed in a combined reaction per sample (plate) for 30 min at 20 °C, followed by thermal enzyme inactivation at 65 °C for 30 min. Adaptor ligation was done using the NEBNext^®^ ligation master mix plus enhancer and the TruSeq unique dual indexed IDT adaptors (Illumina, **Table S2**). Ligation reactions were incubated for 15 min at 20 °C. Ligated sequencing libraries were purified on a Hamilton STAR NGS liquid handler, using two rounds of magnetic SPRI bead purification (0.7X volume).

### ILLUMINA SEQUENCING

Sequencing libraries were pooled (24-36 samples/pool) and quantified using the Qubit dsDNA assay (Thermo Fisher). Samples were diluted appropriately and denatured to a final loading concentration of 10 pM. All samples were sequenced on Illumina MiSeq sequencers using 300-cycle MiSeq v2 reagent kits (Illumina). Each pool was sequenced using dual indexed paired-end sequencing of 150*8*8*150 bp cycles of data acquisition and imaging with a run time of approximately 24 hrs. Basecalling was done in real time using MCS v3.1 and FASTQ files were generated using MiSeq Reporter. At least 15M PF reads (>4.5Gb) with base qualities of >Q30 for at least 90% of bases were collected for each run.

### SEQUENCING DATA ANALYSIS

Amplicon sequences were aligned to the reference genome of the SARS-CoV-2 (NC_045512.2)^2^ using bwa mem^14^, possible PCR duplicates were marked with markduplicates from Picard tools^15^ and reads with less than 50 bases aligned were omitted from the alignment. The resulting aligned filtered reads were used for variant calling with bcftools^14^. For consensus sequence generation only variants reported as homozygous were used. In regions targeted with primers we allowed variants to have allele frequency below one in individual. The consensus sequence was masked with ambiguous nucleotides (N) at positions if the depth of coverage was strictly less than 5 reads after restricting to bases of quality 20 or higher. Consensus sequences with more than 10,000 ambiguous nucleotides were discarded from analysis. The mutations in Table S3 were used to define haplogroups/clades.

For network analysis of haplogroups a median-joining network^8^ of SARS-CoV-2 sequences was generated using data from our sequencing effort in Iceland and from GISAID available on March 22 (Table S3). Only sequences with start positions <=200 and stop positions >=29750 were included in the analysis. For the GISAID sequences, only those with <=1% missing nucleotides were used, whereas for the Icelandic sequences a more permissive threshold of <=5% was imposed. To reduce noise in the network, an imputation step was implemented for sequences with missing nucleotides at sites where other sequences varied, whereby the missing nucelotide was imputed to the consensus variant for the clade (A, A2, A2a, B, B1, B1a, B4, B2, A1a, A3, A6, A7, A2a1, A2a1a, A2a2 or A2a2a) it was assigned to, based on non-missing sites.

Contact tracing information was obtained from the Chief Epidemiologist, Directorate of Health for 577 confirmed cases. The information includes travel abroad, confirmed transmissions, contact with other confirmed cases as well as demographic information. The study was approved by the National Bioethics Committee of Iceland (Approval no. VSN-20-070).

## Results

### RESULTS OF SARS-COV-2 TESTING

The first SARS-CoV-2 infected individual in Iceland was identified on February 28, having returned from northern Italy before it had been designated a high-risk area by the Icelandic authorities. Less than a month later, March 22 2020, 528 individuals had tested positive for the virus out of 4,551 targeted for testing (11.6%) and 50 out of 5,502 participants of the population screening (0.9%) (**Table 1**).

**Table 1.** Description of participants in the population screening and those positive for SARS-CoV-2 in the targeted testing and in the population screening. ^a^One individual traveled to the UK and the Netherlands.

|  | Targeted testing<br>Feb 1-March 15 |  | Population screening<br>March 13-19 |  | Targeted testing<br>March 16-22 |  |
| --- | --- | --- | --- | --- | --- | --- |
|  | All | SARS-CoV-2<br>positive | All | SARS-CoV-2<br>positive | All | SARS-CoV-2<br>positive |
| N | 1,928 | 177 | 5,502 | 50 | 2,623 | 351 |
| Fraction SARS-CoV-2 positive | 10.1% |  | 0.9% |  | 13.4% |  |
| SARS-CoV-2 sequenced | - | 157 | - | 39 | - | 144 |
| N male (%) | 838 (43.5%) | 92 (52.0%) | 2,516 (45.7%) | 29 (58%) | 1,054 (40.2%) | 184 (52.4%) |
| Age (SD) | 40.0 (18.2) | 44.4 (15.7) | 38.2 (18.4) | 42.3 (12.7) | 38.8 (19.0) | 42.0 (15.9) |
| N traveled (%) | - | 115 (65.0%) | 764 (13.9%) | 17 (34.0%) | - | 88 (25.1%) |
| China (high-risk area) | - | 1 | 1 | 0 | - | 0 |
| Austria (high-risk area February 29) | - | 43 | 15 | 0 | - | 29 |
| Italy (high-risk area February 29) | - | 45 | 5 | 0 | - | 2 |
| Switzerland (high-risk area February 29) | - | 9 | 8 | 0 | - | 2 |
| United Kingdom | - | 7 | 155 | 10<sup>a</sup> | - | 20 |
| United States | - | 7 | 122 | 3 | - | 9 |
| Denmark | - | 2 | 81 | 1 | - | 0 |
| Germany (high-risk area March 12) | - | 1 | 27 | 1 | - | 1 |
| Spain (high-risk area March 14) | - | 0 | 112 | 0 | - | 13 |
| Netherlands | - | 0 | 19 | 2<sup>a</sup> | - | 2 |
| Poland | - | 0 | 38 | 1 | - | 0 |
| Other | - | 0 | 386 | 0 | - | 10 |
| Known contact with infected | - | 77 (40.1%) | 125 (2.3%) | 3 (6.0%) | - | 195 (55.6%) |
| Symptoms | - | 153 (86.4%) | 2,441 (44.4%) | 33 (66.0%) | - | 325 (92.6%) |
| Recorded which symptoms | - | 78 | 2,441 | 33 | - | 311 |
| Fever | - | 38 (48.7%) | 393 (7.1%) | 13 (26.0%) | - | 173 (55.6%) |
| Coughing | - | 23 (29.5%) | 1,236 (22.5%) | 21 (42.0%) | - | 110 (35.4%) |
| Body ache | - | 26 (33.3%) | 400 (7.3%) | 13 (26.0%) | - | 138 (44.4%) |
| Headache | - | 21 (26.9%) | 745 (13.6%) | 12 (24.0%) | - | 68 (21.9%) |
| Sore throat | - | 15 (19.2%) | 1,076 (19.7%) | 15 (30.0%) | - | 45 (14.5%) |
| Runny nose | - | 14 (17.4%) | 1,247 (22.8%) | 14 (28.0%) | - | 32 (10.3%) |
| Fatigue | - | 12 (15.4%) | - | - | - | 63 (20.3%) |
| Loss of smell | - | 2 (2.6%) | - | - | - | 21 (6.8%) |
| Other | - | 53 (23.5%) | 603 (11.0%) | 9 (18.0%) | - | 42 (13.5%) |

Sample collection for population screening started March 13 and the first positive test results were communicated to the Icelandic health authorities on March 15. Because of this and the rapid changes in definition of high-risk areas by the Icelandic authorities, we divide the targeted testing into early testing (Feb 1-March 15) and later testing (March 16-22). In the early targeted testing, 65.0% of those that tested positive for SARS-CoV-2 (positives) had recently traveled outside Iceland, while in the later targeted testing 25.1% had (**Table 1**). In the early targeted testing 40.1% of positives reported contact with a known infected individual, compared to 55.6% in the later. Of the positives from the population screening, 34.0% had recently traveled outside of Iceland, which is substantially more than the 13.7% of the negatives (P=0.00025), and only 6% of them reported contact with an infected person. This low number of contacts is most certainly because known contacts of infected persons were in quarantine and were not eligible for the population screening.

Strikingly, among the positives from the early targeted screening who had traveled, 86.1% had traveled to areas that were designated as high-risk areas by the end of February (China and the Alp regions in Austria, Italy, and Switzerland), while none of the positive individuals from the population screening had traveled there, probably because of the effective quarantine of individuals arriving from these regions. On the other hand, a disproportionately high fraction of positives from the population screening (20.0%) had traveled to the UK, compared to the negatives (2.8%, P=9.2e-7), suggesting relatively early spread of the virus in the UK population.

Eighty six percent of positives from the targeted testing reported symptoms whereas sixty six percent of those positive from the population screening reported symptoms. However, 44.2% of negative individuals also reported symptoms. Fever and body aches were most enriched among positive vs negative individuals in the population screening. Fever, coughing, and body aches were the most common symptoms among the positive in the targeted testing.

The individuals targeted for testing were of similar ages (mean=39.3, SD=18.7) as the participants of the population screening (mean=38.4, SD=18.4) (**Figure 1**). For both sets, the positives are older and have a narrower age distribution (**Table S1**). Specifically, out of the 354 and 433 individuals under the age of 10 only 10 (2.8%) and 0 (0.0%) tested positive in the targeted testing and population screening groups, respectively. This is substantially lower than the fraction of 12.3% (P=1.6e-9) and 1.0% (P=0.031) among the 4,551 and 5,502 individuals over 10 years of age in the targeted testing and population screening, respectively. Only 61 individuals over 80 years of age were tested in the targeted testing and population screening combined and only one of these individuals was positive, less than would be expected from the fraction of positives below 80 years of age (P=0.0080).

**Figure 1.**
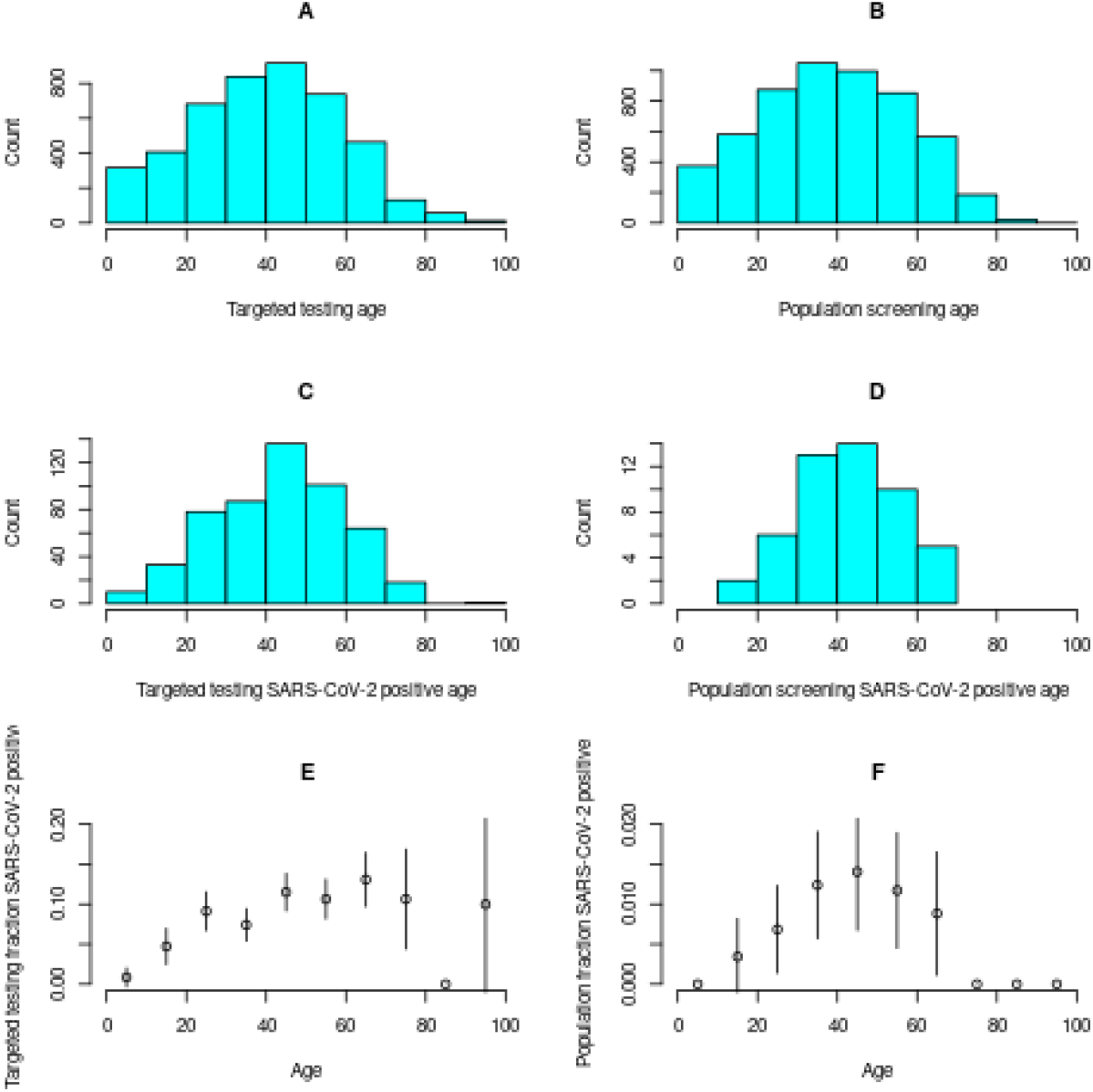
The age distribution of those targeted for testing and participating in population screening, the age distribution of the ones that are SARS-CoV-2 positive, and the fraction of positive by 10 year age bins. The age distribution of individuals targeted for testing and participating in the population screening are shown in panels A and B. The age distribution of the subset that tested positive are shown in panels C and D. The fraction of individuals that tested positive is shown in panels E and F.

In both the targeted testing and population screening, more females were tested than males or 57.4% and 54.3%, respectively (**Table 1**). However, the fraction of males that tested positive was greater than the fraction of females: 14.6% vs 9.5% in the targeted testing were positive (P=6.8e-9 after adjusting for age) and 1.1% vs 0.7% in the population screening (P=0.070 after adjusting for age, **Figure 2**).

**Figure 2.**
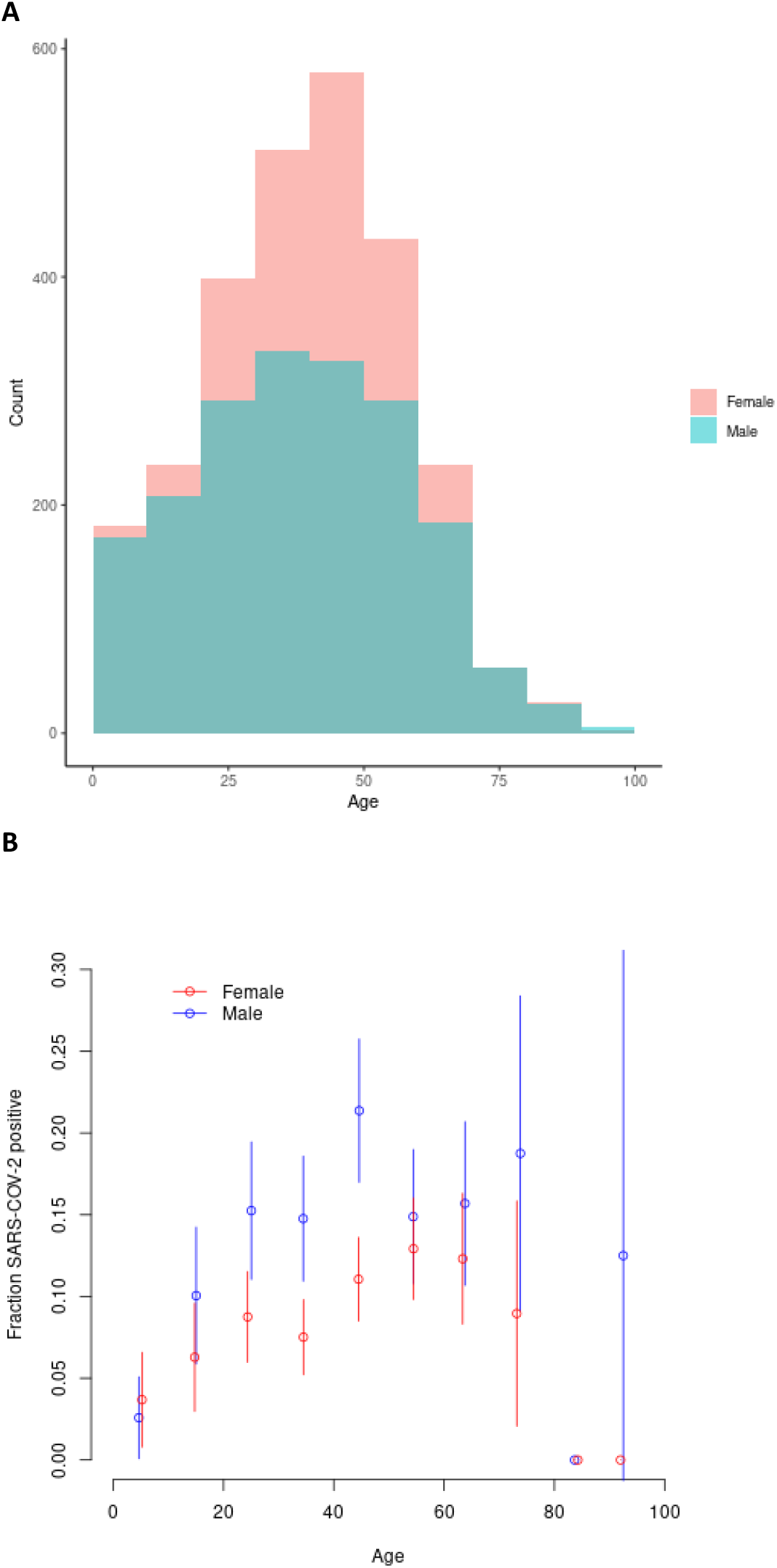
A) Age distribution of those targeted for testing by sex and B) the fraction of SARS-CoV-2 positive by age and sex.

### HAPLOTYPES OF SARS-COV-2

We sequenced RNA extracted from 378 positives and managed to cover more than 90% of the SARS-CoV-2 genome from 343 samples and 67% from 355 samples. We called 251 sequence variants, 128 of which were not found in the GISAID database (**Figure 3A**). We used clade informative mutations (**Table S3**) to assign 340 individuals to haplogroups: 301 identified from targeted testing and 39 from population screening (**Table 2, Figure 3**).

**Table 2.** The distribution of SARS-CoV-2 haplotypes by time and transmission mode. Also shown are the countries traveled to before being tested positive. Countries are represented by two-letter country codes: AT:Austria, CH:Switzerland, DE:Germany, DK:Denmark, ES:Spain, IT:Italy, NL:Netherlands, UK: United Kingdom, US:USA.

| Hap | Early targeted testing<br>Feb 1-March 15 |  |  |  |  |  | Population screening<br>March 14-19 |  |  |  |  |  | Later targeted testing<br>March 16-22 |  |  |  |  |
| --- | --- | --- | --- | --- | --- | --- | --- | --- | --- | --- | --- | --- | --- | --- | --- | --- | --- |
|  | Imported |  |  | Local |  |  | Imported |  |  | Local |  |  | Imported |  |  | Local |  |
|  | N | % | Country | N | % |  | N | % | Country | N | % |  | N | % | Country | N | % |
| A1a | 2 | 2.0 | IT:1, CH:1 | 6 | 10.7 |  | 4 | 26.7 | UK:4 | 14 | 58.3 |  | 3 | 7.5 | ES:2, UK:1 | 18 | 17.3 |
| A2a1 | 36 | 35.6 | IT:29, AT:3 | 16 | 28.6 |  | 4 | 26.7 | UK:3, DE:1 | 3 | 12.5 |  | 7 | 17.5 | AT:4, UK:2 | 15 | 14.4 |
| A2a | 24 | 23.8 | IT:11, AT:6 | 16 | 28.6 |  | 2 | 13.3 | NL:1, DK:1 | 7 | 29.2 |  | 19 | 47.5 | AT:12, UK:3 | 47 | 45.2 |
| B1a | 4 | 4.0 | US:4 | 0 | 0 |  | 1 | 6.7 | US:1 | 0 | 0.0 |  | 5 | 12.5 | US:5 | 2 | 1.9 |
| A2a2 | 33 | 32.7 | AT:28, DK:2 | 18 | 32.1 |  | 2 | 13.3 | US:2 | 0 | 0.0 |  | 5 | 12.5 | AT:4, ES:1 | 17 | 16.3 |
| A8 | 2 | 2.0 | UK:1, AT:1 | 0 | 0 |  | 0 | 0.0 |  | 0 | 0.0 |  | 0 | 0.0 |  | 0 | 0.0 |
| A10 | 0 | 0.0 |  | 0 | 0 |  | 0 | 0.0 |  | 0 | 0.0 |  | 1 | 2.5 | UK:1 | 1 | 1.0 |
| A9 | 0 | 0.0 |  | 0 | 0 |  | 2 | 13.3 | UK:2 | 0 | 0.0 |  | 0 | 0.0 |  | 4 | 3.8 |

**Figure 3.**
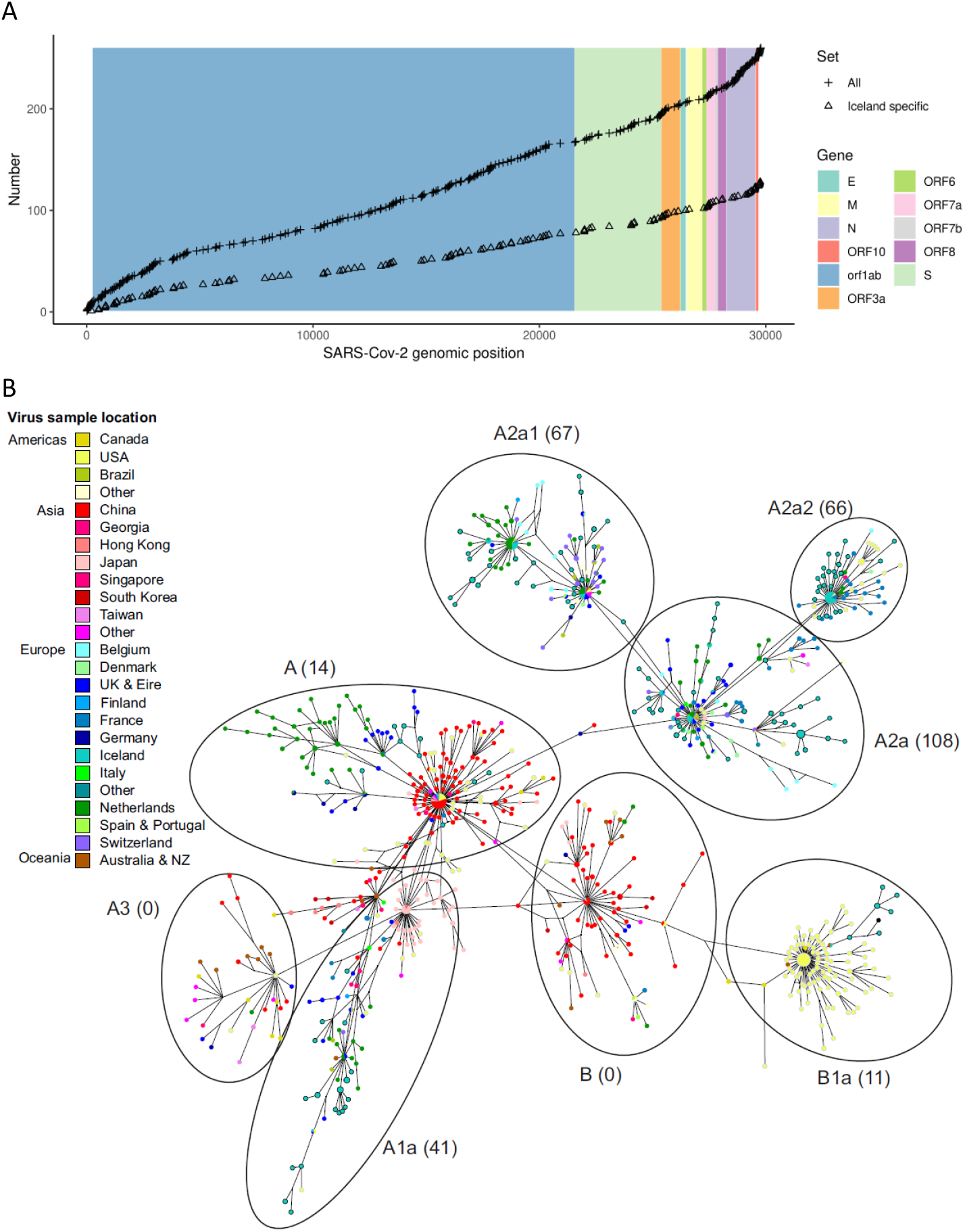
A) The distribution of variants across the SARS-CoV-2 genome B) A median-joining network of 1,341 SARS-CoV-2 sequences. Each circle represents a different sequence type, where circle size reflects the number of carrier hosts and lines between circles represent one or more mutations that differentiate the sequence types. Circles are colored by the countries or regions where hosts were sampled. The principal clades are delineated by labeled elipses with the number of sequences from Icelanders shown in parentheses.

To shed further light on the geographical origin of the SARS-CoV-2 infections in Icelanders, we generated a median-joining network of 1,341 complete virus sequences (307 from complete genomes from Icelanders and 1,034 from other populations around the world). Figure 3B shows that several virus lineages (clades) have emerged during the 3-4 months since the original outbreak in China, with an average of 5 mutations separating them from the founding haplotype from Wuhan (the central haplotype of clade A). Although the sequencing efforts vary considerably among populations (there is for example an underrepresentation from Italy and an overrepresentation from Iceland and the Netherlands), it is evident that the geographical distribution of clades is highly structured. Thus, clades A and B are common in East Asia, whereas clade B1a appears to be at the center of the outbreak on the West coast of the United States and clade A2a and its descendants are almost exclusively found in European populations.

Of the targeted testing positives, 157 were sampled in early testing and the remaining 144 in later testing. The SARS-CoV-2 infections observed in Iceland cluster into eight clades, seven A clades and one B clade.

Most haplotypes from the early targeted testing were of the A2 clade (143 out of 157 haplotypes) (**Figure 4**). By the time we initiated the population screening, all travelers returning from ski resorts in the Alps were requested to self-quarantine and were not eligible for participating, which resulted in a substantially different composition of haplotypes. For example, the A2a2 haplotype which was most commonly seen in travelers coming from Austria in the early targeted testing was much rarer in travelers in the population screening and completely absent from those who had not traveled. Interestingly, the A1a haplotype was more common in the population screening than the targeted testing with a total of 18 out of 39 haplotypes in the population screening compared to only 8 out of 157 haplotypes in the early targeted testing. It was one of the most common haplotypes in travelers in the population screen and it was by far the most common haplotypes seen in those who had not traveled.

**Figure 4.**
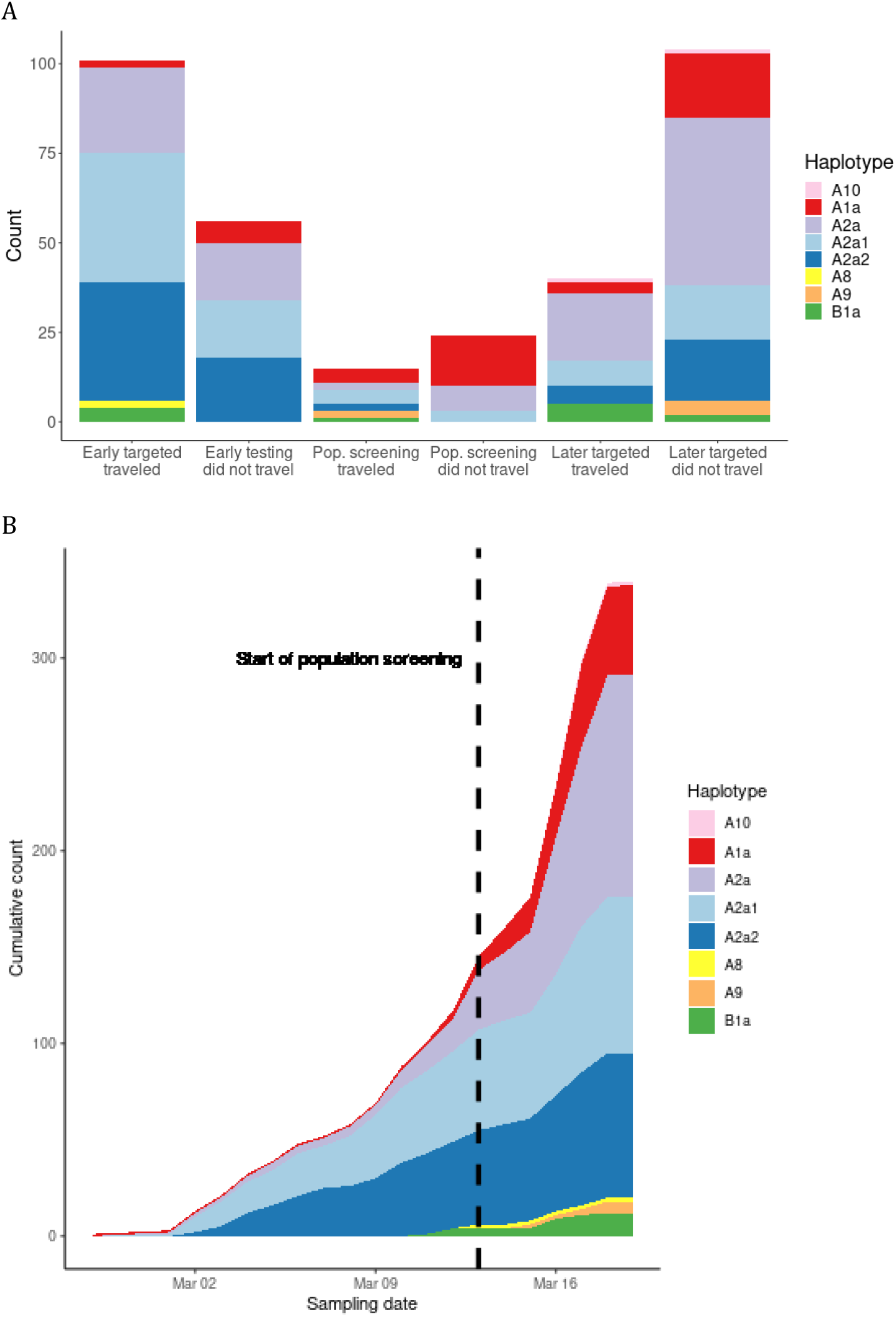
The distribution of SARS-CoV-2 haplotypes depending on sampling and travel status. A) The counts of each of the eight haplotypes seen in Iceland is shown depending on whether the positive was found through early targeted testing, population screening, or later targeted testing and whether the positive had recently traveled outside Iceland. B) The cumulative haplotype counts from targeted testing and population screening are shown as a function of sampling date. The dashed vertical line indicates the start of the population screening.

The composition of haplotypes changed substantially from early targeted testing to later targeted testing (**Figure 4**). The A2a1 and A2a2 haplotypes which had collectively made up 103 out of 157 haplotypes (65.6%) in the early targeted testing were reduced to 44 of 144 haplotypes (30.6%) in the late targeted testing, mostly because of the increased frequency of the A1a and A2a haplotypes, the two most common haplotypes in the population screening. This is probably because the population screening identified clusters of infected individuals who seeded from areas that had not been designated as high-risk e.g. the UK. The later targeted testing was then more likely to find new individuals from these clusters, both because the targeted testing was extended to include more high-risk areas and because the population screening had identified cases that could be used to inform further contact tracking efforts.

### HAPLOTYPE ANALYSIS OF CONTACT TRACING NETWORKS

Haplotype analysis based on SARS-CoV-2 sequences overlaid on contact tracing networks show good concordance between the contacts identified by the tracing team and those based on viral sequences (**Figure 5A**). Of the 219 pairs of individuals found through contact tracing, 183 were consistent with the sequencing data.

**Figure 5.**
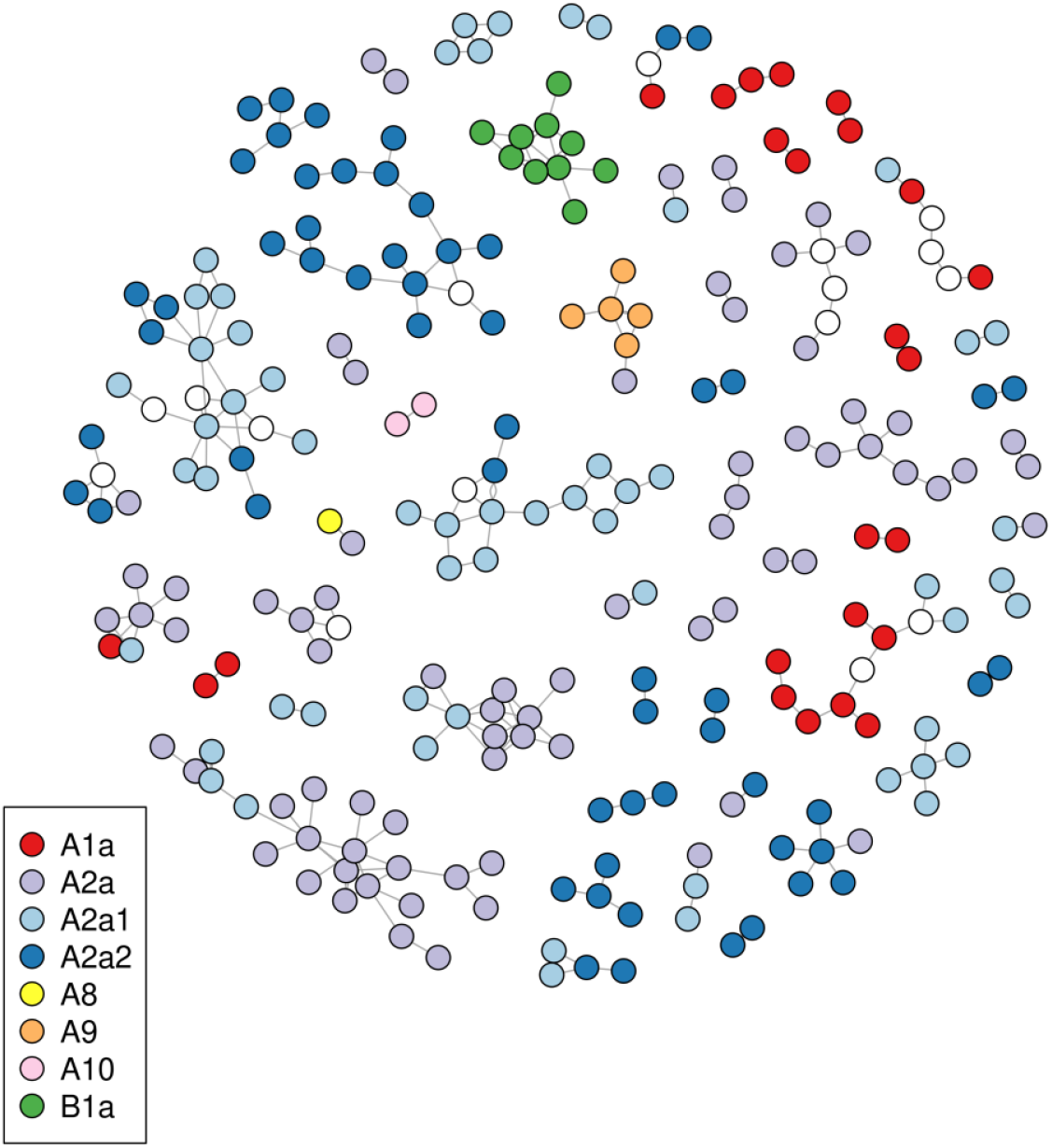
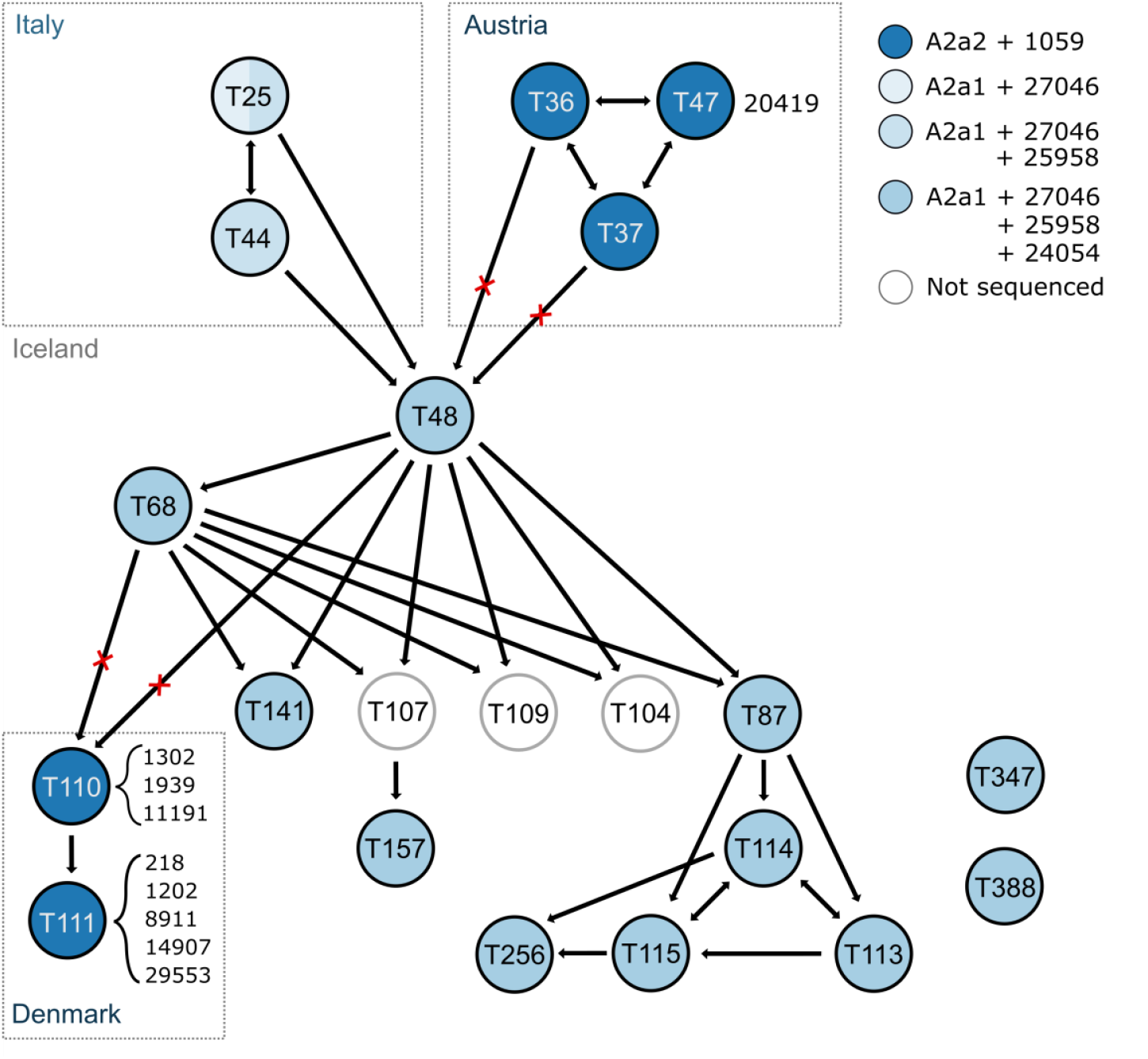
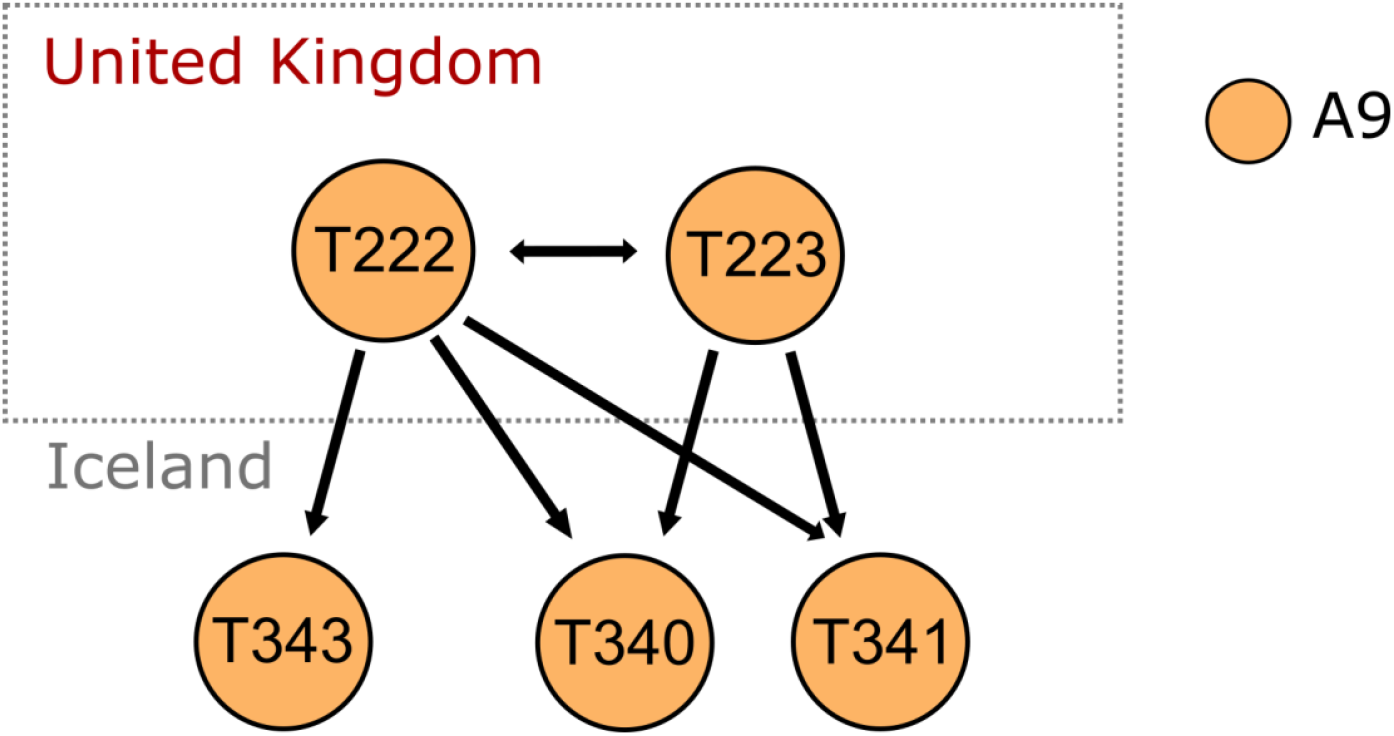
(A) An overview of all clusters in the contact tracing network with sequenced viruses (B) A contact tracing network for cluster including a novel domestic mutation (24054C>T). T25 carries both the A2a1a strain and the A2a1a+25958 strain. (C) A contact tracing network for individuals tested positive through population screening at deCODE. Contact tracing networks show infected individuals as nodes and a connection between two nodes where a transmission of infection or contact has been established. In cases where the direction of transmission is ambiguous a bidirectional arrow is displayed. Individuals that traveled internationally are drawn in boxes representing their travel destination. The colors of nodes represent the haplotype of the viral strain, either as a clade or a clade plus one or more mutations. The labels on the nodes are identifiers given in increasing order of identification, e.g. T6 is the 6th case reported. Additional mutations are represented by position number besides each node. Red x marks indicate recorded contacts that are inconsistent with the viral haplotypes carried by the individuals.

**Figure 5B** shows one of the most complex contact tracing networks, where clusters of individuals returning from Italy or Austria connect with individuals infected in Iceland. The figure shows a network of 14 individuals who were infected in Iceland and their infections could be traced to two possible sources in Austria and northern Italy through individual T48. Haplotype analysis showed that these individuals were infected by viruses with the A2a1 haplotype, which has more commonly been imported from northern Italy than Austria (**Table 2**). Supporting this route of transmission, we find novel mutations on the A2a1 background (24054C>T and 25958A>G) specific to this cluster, present in all viral sequences from individuals downstream of T48. Interestingly, one of the individuals coming from northern Italy (T25) carries both the 25958 mutation and the wild type, so that only a subset of the virus infection of T25 has the mutation, whereas only the 25958 is carried by the other (T44). Neither carry the 24054 mutation. However, all downstream individuals carry both mutations. This suggests that the 24054 mutation is a novel domestic mutation, whereas the 25958 mutation was likely present in northern Italy, although it is not found in the Nextstrain public database.

Additionally, two individuals were also associated with this cluster (T110 and T111). However, they carry a distinct strain from the rest of the cluster, potentially acquired from travel to Denmark.

The rare mutations (24054C>T and 25958A>G) that we identified in the cluster outlined in **Figure 5B** are highly informative about domestic spread in Iceland. We searched for individuals carrying these mutations who were not reported to associate with this cluster. This resulted in two individuals who had probably been infected by someone in the cluster through an unknown contact: T347 who tested positive in targeted testing and T388 discovered through the population screening.

We informed the tracking team about positive individuals in the population screening. They then through tracking identified other positive individuals whom they had been in contact with. For example, the top two individuals in Figure 5C tested positive in the population screening with a distinct A clade (A9) after travel to the UK. Their contacts were traced and three later tested positive through targeted testing with the A9 clade. Overall, our extensive genomic tracking adds resolution of tracing transmission routes.

## Discussion

In this paper we describe how SARS-CoV-2 has entered and spread through the Icelandic population. The conditions in Iceland are favorable for a study of this kind. There is only one major gateway into the country and a single payer healthcare system with universal access. SARS-CoV-2 infected individuals were identified both through targeted testing of individuals at high risk of infection and by population screening.

As of March 19, 5,502 individuals had participated in the population screening and of those 50 tested positive for the virus (0.9%). There are weaknesses in the design of this screening: the sample is not random as all residents of Iceland were invited to participate and those concerned about potential infection are more likely to participate than others. We asked people with symptoms of COVID-19 not to participate but to seek help in the healthcare system. However, close to half of participants reported symptoms, most commonly a rhinorrhea and coughing.

The healthcare system began testing of individuals considered to be at risk of infection on January 31 2020 and tested 65 before the first infected was identified on February 28. By March 22, 4,511 had been tested and 528 (11.6%) were positive. Hence those at high risk of infection were eleven times more likely to test positively.

We show here that young children and females are less likely to test positive for SARS-CoV-2 than adults and males, respectively. Others have shown that children and females are less likely to develop severe disease than adults and males, respectively^7,8^. These results are consistent and suggest that children and females are less vulnerable to SARS-CoV-2.

To further our understanding of the spread of the virus and to determine how the virus mutates as it spreads, we sequenced the virus from 357 individuals who had tested positive before March 19. The haplotype composition of the viruses from individuals identified through the population screening was different from those tested in the early targeted testing; more had the A1a haplotype and fewer had the A2a haplotypes. Hence, it looks like the haplotypes of the virus that are propagating in the general population come from a different source, perhaps brought in by people coming from countries that had not yet been designated high-risk areas. Furthermore, the sequence diversity in the virus from the population screening is greater than in the original targeted screening that was focused on people coming home from skiing in the Alps. Hence, it is likely that the virus has been carried into Iceland from many countries.

Since the first individual tested positive for SARS-CoV-2 in Iceland on February 28, a specialized team has identified contacts of positives and they have been instructed to self-quarantine for 14 days. On March 22, 6,816 were in quarantine and 1,193 had completed quarantine. Most of the individuals who test positive by targeted testing were already in quarantine. This supports wisdom of the actions of the Icelandic health care authorities; namely targeted testing, isolation of the infected and tracking down contacts for quarantine. Further, authorities have emphasized public education regarding hygiene measures and social distancing as well as implementing a public gathering ban. Several measures have also been incorporated to protect the elderly and other groups are at greater risk of serious illness should they develop COVID-19.

The 0.9% frequency of the infection in the population screening indicates that the virus is spreading to the extent that unless we increase the screening effort we are likely to fail in our efforts to contain it.

## Data Availability

All sequenced virus genomes have been deposited into GISAID.

